# Expert Opinion on COVID-19 Vaccination and the Use of Cladribine Tablets in Clinical Practice

**DOI:** 10.1101/2021.06.22.21259308

**Authors:** Peter Rieckmann, Diego Centonze, Gavin Giovannoni, Le H. Hua, Celia Oreja-Guevara, Daniel Selchen, Per Soelberg Sørensen, Patrick Vermersch, Heinz Wiendl, Hashem Salloukh, Bassem Yamout

**Author notes:** **Corresponding author:** Bassem Yamout. Neurology Institute, Harley Street Medical Center, Abu Dhabi, UAE.

## Abstract

**Background:** Gaps in current evidence and guidance leave clinicians with unanswered questions on the use of cladribine tablets for the treatment of multiple sclerosis (MS) in the era of the COVID-19 pandemic, in particular relating to COVID-19 vaccination.

**Objective:** We describe a consensus-based program led by international MS experts with the aim of supplementing current guidelines and treatment labels by providing timely recommendations relating to COVID-19 vaccination and the use of cladribine tablets in clinical practice.

**Methods:** A steering committee (SC) of 10 international MS experts identified seven clinical questions to answer concerning the use of cladribine tablets and COVID-19 vaccination, which addressed issues relating to patient selection, timing and efficacy, and safety. Clinical recommendations to address each question were drafted using available evidence combined with expert opinion from the SC. An extended faculty of 28 MS experts, representing 19 countries, in addition to the 10 SC members, voted on the recommendations. Consensus on recommendations was achieved when ≥75% of respondents expressed an agreement score of 7–9, on a 9-point scale.

**Results:** Consensus was achieved on all 13 recommendations. Clinical recommendations are provided on whether all patients with MS receiving cladribine tablets should be vaccinated against COVID-19, and whether they should be prioritized; the timing of vaccination around dosing of cladribine tablets (i.e., before and after a treatment course); and the safety of COVID-19 vaccination for these patients.

**Conclusions:** These expert recommendations provide timely guidance on COVID-19 vaccination in patients receiving cladribine tablets, which is relevant to everyday clinical practice.

## Introduction

The COVID-19 pandemic has represented a global public-health emergency, which has affected both acute and ongoing medical care. Multiple sclerosis (MS) is a lifelong, progressive disease that requires regular management and treatment. MS, including the relapsing forms of MS, in which the patient experiences periods of stability in between episodes of new or worsening symptoms, is typically treated by disease-modifying therapies (DMTs) such as cladribine tablets (Mavenclad^®^; Merck Europe B.V., The Netherlands). Many DMTs, including cladribine tablets, affect immune response, and this is the rationale used to treat an autoimmune disease such as MS. The effect of DMTs on immune cells may increase the risk of infections as well as impair vaccine response. The provision of health and social-care services for MS have been significantly affected by the COVID-19 pandemic, leading to treatment delays and interruptions to rehabilitation services. In some parts of the world this has affected the health and wellbeing of people with MS, including the risk of disease progression.^1-4^ At the time of writing, there have not been any efficacious therapies for the treatment of SARS-CoV-2 infection.^5^

Vaccination against COVID-19 remains the most efficient way to protect the global population, including people with MS. Approved vaccines have demonstrated high effectiveness and acceptable safety, yet hesitation to be vaccinated against COVID-19 remains a challenge in some people with MS.^6, 7^ All DMTs used in the treatment of MS lack vaccine-specific studies for SARS-CoV-2 due to its novel nature, although vaccine studies exist for other pathogens. This raises practical difficulties related to timing of vaccination relative to MS treatment, given treatment modulation of the immune system. Stopping or delaying treatment can potentially result in disease rebound. In addition, and given that MS is an autoimmune disease, there is always a theoretical concern for the effect of vaccination on MS disease status.

Cladribine tablets are a short-course, oral DMT for use in MS, approved by the European Medicines Agency and the United States Food and Drug Administration (FDA), as well as other regulatory authorities around the world.^8-11^ Cladribine is a deoxyadenosine analogue that selectively reduces B and T lymphocytes and is thought to interrupt the cascade of immune events central to the pathogenesis of MS.^9^ Common adverse reactions reported in MS patients who receive cladribine tablets at the recommended cumulative dose of 3.5 mg/kg body weight over 2 years in clinical studies included oral herpes, herpes zoster, rash, alopecia, and decrease in neutrophils count, while lymphopenia was very common. Local and global medical societies have issued national and international guidance on the use of DMTs for MS during the COVID-19 pandemic, and on COVID-19 vaccination in patients receiving DMTs.^12-17^ These guidelines and position statements, however, leave gaps and unanswered questions, and more specific guidance for everyday clinical practice is required; these guidelines and position statements are summarized in Supplementary Table S1.

To improve the care provided to people with MS treated with cladribine tablets during the ongoing global pandemic, a consensus-based program was developed to address unanswered questions relating to patient selection for COVID-19 vaccination, vaccine timing, efficacy, and safety. The objective of the program was to provide timely, consensus-based, practical recommendations relating to COVID-19 vaccination in MS patients receiving cladribine tablets.

## Materials and methods

The consensus program, based on a modified Delphi methodology, took place between February and May 2021; the process is outlined in Figure 1. The methodology was used previously for a consensus paper on the use of cladribine tablets in clinical practice.^18^ Merck KGaA (Darmstadt, Germany) provided funding for the project, but had no input into the development of clinical questions nor recommendations. No company representative voted on the recommendations. A steering committee (SC) of 10 international MS experts led the program, co-chaired by Peter Rieckmann and Bassem Yamout. The SC developed the following seven clinical questions to be addressed with the consensus:

**Figure 1:**
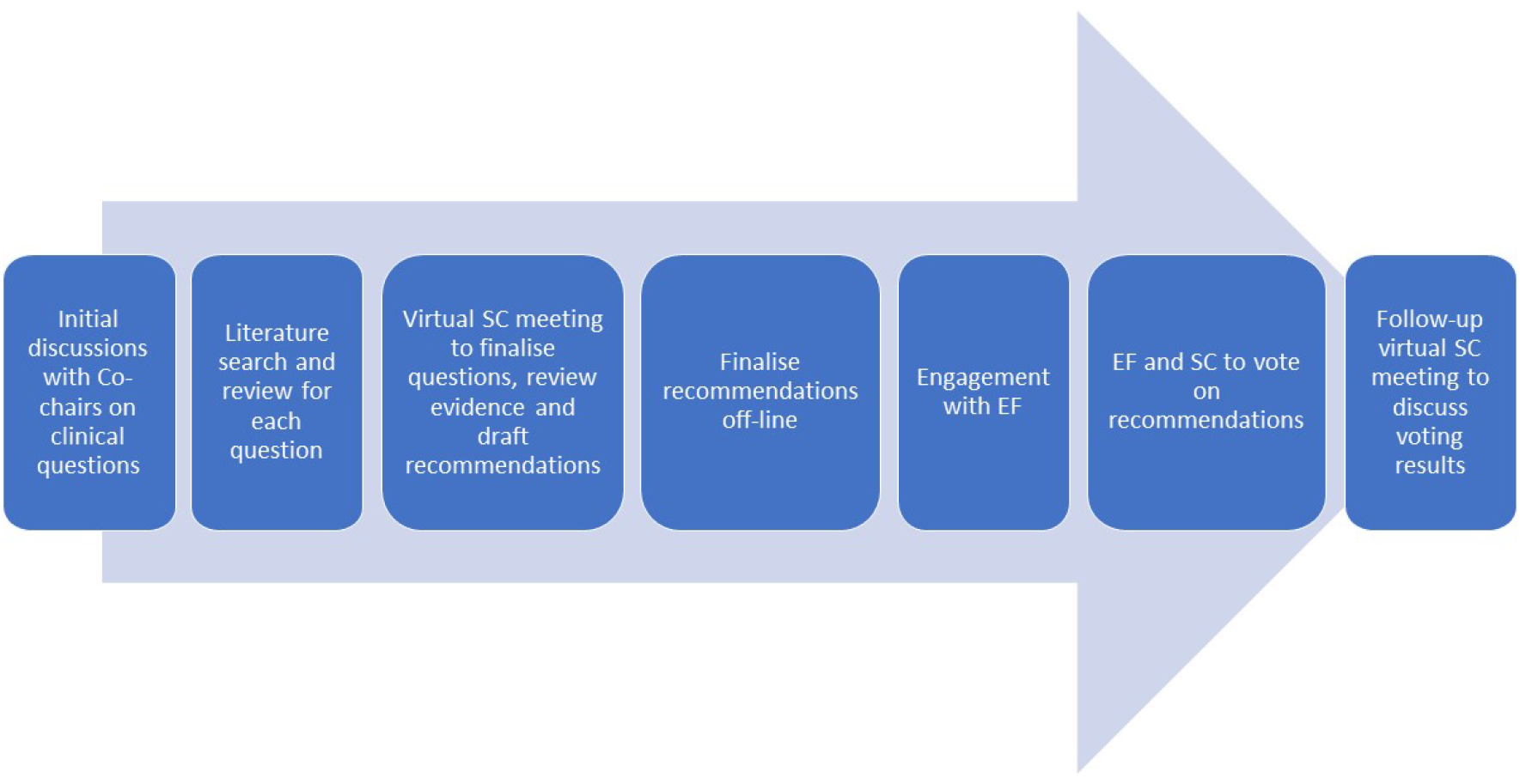
Schematic of the modified Delphi process for achieving consensus. EF, extended faculty; SC, steering committee

1. Should all people with MS receiving cladribine tablets be vaccinated against COVID-19?
2. Should people with MS treated with cladribine tablets be prioritized for COVID-19 vaccination?
3. Should all people with MS on cladribine tablets receive a vaccination against COVID-19 at the time it is offered?
4. When should a person with MS be vaccinated against COVID-19 if they are about to start treatment with cladribine tablets (first course in Year 1 or second course in Year 2)?
5. When should a person with MS be vaccinated against COVID-19 if they are already undergoing treatment with cladribine tablets (post-course one, or post-course two)?
6. Based on currently available data, are the COVID-19 vaccines safe for use in people with MS receiving cladribine tablets?
7. Will COVID-19 vaccination lead to exacerbation of MS symptoms or relapse while undergoing treatment with cladribine tablets?

A comprehensive literature review was performed using the PICO (Population, Intervention, Comparison, Outcome) framework for each of the seven questions. The level of evidence was assessed and agreed by the SC using the GRADE (Grading of Recommendations Assessment, Development, and Evaluation) level of evidence ratings scale.^19^ After reviewing the available evidence, the SC drafted clinical recommendations to answer the questions.

An extended faculty (EF) of MS experts were invited to vote on the clinical recommendations. A large number of experts were invited to take part in the project as EF, with the aim of achieving an approximate participation rate of 25%. A total of 82 experts from 27 countries were invited. The final EF comprised 28 international experts from 19 countries (a participation rate of 34%). Clinical recommendations were voted on by the SC and EF members (N=38). Consensus was achieved when ≥75% of respondents agreed in the range 7–9 (on a 9-point scale). Each statement/recommendation was assigned a strength score (i.e., the median score) and a level of consensus, defined as the percentage of votes with a score of 7–9.^18, 20-24^

## Results

In total, 13 recommendations were drafted by the SC for voting. Consensus was achieved on all 13 of these recommendations, with 12 recommendations achieving consensus in the range of 90–100% and one recommendation achieving consensus in the range of 80–90%. All but two EF members voted on all questions. These abstentions were due to technical issues with the online platform. Therefore, the number of experts voting on each recommendation was either 36 or 38. A summary of the available evidence for each question is given below, and the clinical recommendations for each question provided in Tables 1–3. For those recommendations where some faculty members voted 6 or less, their reasons for doing so are listed in Supplementary Tables S2–S4.

**Table 1.**
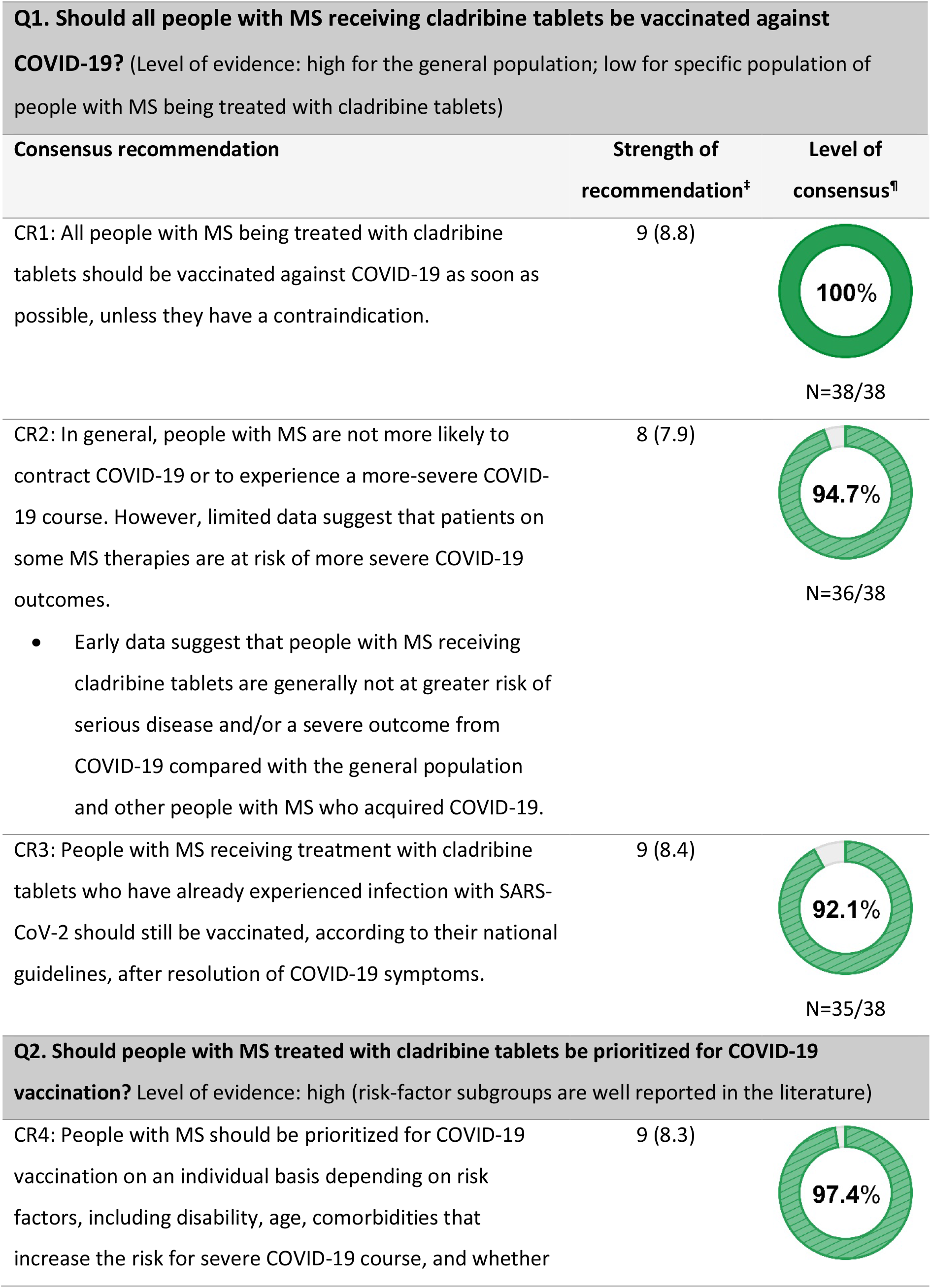
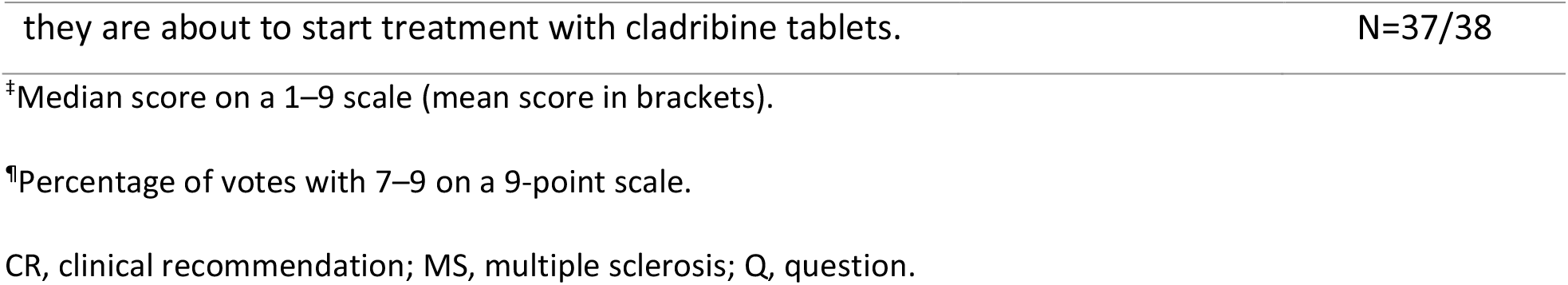
Consensus recommendations to address clinical questions on patient selection for COVID-19 vaccination.

| **Q1. Should all people with MS receiving cladribine tablets be vaccinated against COVID-19?** (Level of evidence: high for the general population; low for specific population of people with MS being treated with cladribine tablets) |  |  |
| --- | --- | --- |
| Consensus recommendation | Strength of recommendation<sup>‡</sup> | Level of consensus<sup>¶</sup> |
| CR1: All people with MS being treated with cladribine tablets should be vaccinated against COVID-19 as soon as possible, unless they have a contraindication. | 9 (8.8) | 100%<br>N=38/38 |
| CR2: In general, people with MS are not more likely to contract COVID-19 or to experience a more-severe COVID-19 course. However, limited data suggest that patients on some MS therapies are at risk of more severe COVID-19 outcomes.<br>• Early data suggest that people with MS receiving cladribine tablets are generally not at greater risk of serious disease and/or a severe outcome from COVID-19 compared with the general population and other people with MS who acquired COVID-19. | 8 (7.9) | 94.7%<br>N=36/38 |
| CR3: People with MS receiving treatment with cladribine tablets who have already experienced infection with SARS-CoV-2 should still be vaccinated, according to their national guidelines, after resolution of COVID-19 symptoms. | 9 (8.4) | 92.1%<br>N=35/38 |
| **Q2. Should people with MS treated with cladribine tablets be prioritized for COVID-19 vaccination?** Level of evidence: high (risk-factor subgroups are well reported in the literature) |  |  |
| CR4: People with MS should be prioritized for COVID-19 vaccination on an individual basis depending on risk factors, including disability, age, comorbidities that increase the risk for severe COVID-19 course, and whether | 9 (8.3) | 97.4% |

<sup>‡</sup>Median score on a 1–9 scale (mean score in brackets).
<sup>¶</sup>Percentage of votes with 7–9 on a 9-point scale.
CR, clinical recommendation; MS, multiple sclerosis; Q, question.

### Patient selection

#### Question 1. Should all people with MS receiving cladribine tablets be vaccinated against COVID-19?

There is a high level of evidence in the literature for the efficacy of COVID-19 vaccines in the general population. In Phase 3 randomized controlled trials, vaccines (ChAdOx1 nCoV-19, Gam-COVID-Vac (Sputnik V), mRNA-1273, and BNT162b2) demonstrated between 62% and 95% efficacy at preventing COVID-19 illness.^25-28^ Furthermore, real-world evidence of nationwide mass vaccination in Israel suggests that the BNT162b2 mRNA vaccine is effective for a wide range of COVID-19–related outcomes, including symptomatic COVID-19, hospitalization, severe disease, and death.^6^ While the majority of people with MS are not thought to be at a greater risk of COVID-19 disease than the general population, certain exceptions to this exist. In general, older age, male sex, higher Expanded Disability Status Scale (EDSS), longer MS duration, presence of comorbidities, progressive MS course, recent methylprednisolone use, or therapy with an anti-CD20 agent were all risk factors for a more severe COVID-19 disease course.^29, 30^ Similar findings were also observed in a large cohort analysis of 12 data-sources from 28 countries, (n=2340 patients).^31^ Limited data are available on outcomes from SARS-CoV-2–infected MS patients on cladribine tablets. However, analysis of the first 272 cases of COVID-19 (confirmed or suspected) reported to the Merck Global Patient Safety Database (as of 15 January 2021) indicate that people with MS receiving cladribine tablets are generally not at greater risk of serious disease and/or a more-severe outcome with COVID-19 compared with the general population and other patients with MS who acquired COVID-19.^32^ Clinical recommendations to address question 1 are provided in Table 1.

#### Question 2. Should people with MS treated with cladribine tablets be prioritized for COVID-19 vaccination?

Data from a small number of people with MS receiving cladribine tablets indicated that they are not generally at greater risk of serious COVID-19 disease and/or a more-severe outcome.^32^ High-risk subgroups of people with MS have been reported in the literature, which may affect risk, regardless of DMTs. Age, EDSS, and obesity were independent risk factors for severe COVID-19 disease in a multicenter, retrospective, observational cohort study investigating clinical outcomes in 347 patients.^33^ People with MS not on DMTs, with previous cardiovascular diseases, or with a severe degree of disability were identified as more at risk of severe COVID-19 disease in a review of 873 cases of SARS-CoV-2 infection in people with MS.^34^ The clinical recommendation to address question 2 is provided in Table 1.

### Timing and efficacy

#### Question 3: Should all people with MS on cladribine tablets receive a vaccination against COVID-19 at the time it is offered?

While there are no controlled clinical studies specifically investigating vaccine responses in people with MS receiving cladribine tablets, there are data from a small number of patients in the post-approval MAGNIFY-MS and CLOCK-MS trials.^35, 36^ In the MAGNIFY-MS study (NCT03364036), some patients received vaccinations during the trial as standard of care; this presented an opportunity to investigate vaccine responses. Retrospective analysis (N=15) demonstrated that seroprotective antibody levels against varicella zoster and seasonal influenza were maintained or increased for at least 6 months (time of follow-up) independent of lymphocyte counts and irrespective of vaccine timing relative to dosing with cladribine tablets.^36^ In the ongoing CLOCK-MS study (NCT03963375), a small subset of enrolled patients who had received treatment with cladribine tablets and planned to receive the influenza vaccine as part of standard care were enrolled in the vaccine substudy.^35^ Initial findings (N=4) show seroprotective influenza antibody levels at four weeks post-vaccination in MS patients taking cladribine tablets, independent of lymphocyte counts (including two patients—one with grade 1 and one with grade 2 lymphopenia—at the time of vaccination).^35^ Recently updated findings from 272 reported cases of COVID-19 among cladribine tablets recipients found no greater risk of serious and/or severe outcomes compared to the general population, with a total of 15% (40/272) of patients experiencing a severe course of COVID-19.^32^ Data on immune cell responses following treatment with cladribine tablets may have relevance to the timing of vaccinations. Cladribine tablets preferentially reduce cells of the adaptive immune system, while leaving the innate immune system relatively spared. There is a preferential reduction in lymphocyte subpopulations, followed by lymphocyte count recovery (immune reconstitution). The lowest absolute lymphocyte counts occurred approximately 2–3 months after the start of each treatment course and were lower with each additional treatment course. The majority of cladribine tablet–treated patients in the Phase 3 trials experienced grade 1 or 2 lymphopenia; grade 3 lymphopenia was observed in approximately 25% of patients, and <1% of patients experienced grade 4 lymphopenia at any time over 2 years of treatment. In patients receiving cladribine tablets, no increase in opportunistic infections was shown.^37^ Recent data from the MAGNIFY-MS biomarker study showed rapid repopulation of naïve B-cells and the sparing of immunoglobulin G up to 1 year after cladribine tablet treatment.^36^ Clinical recommendations to address question 3 are provided in Table 2.

**Table 2.**
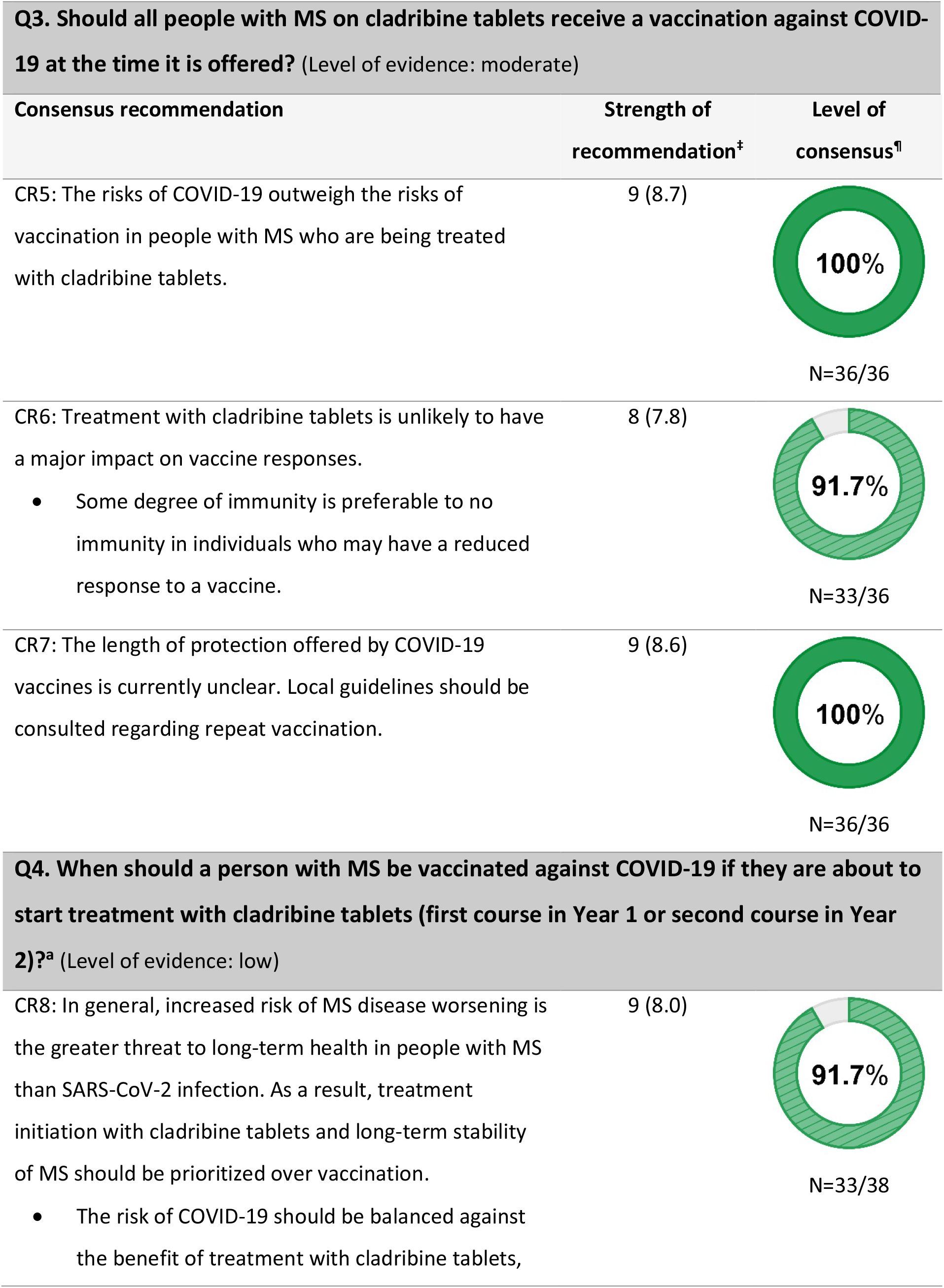
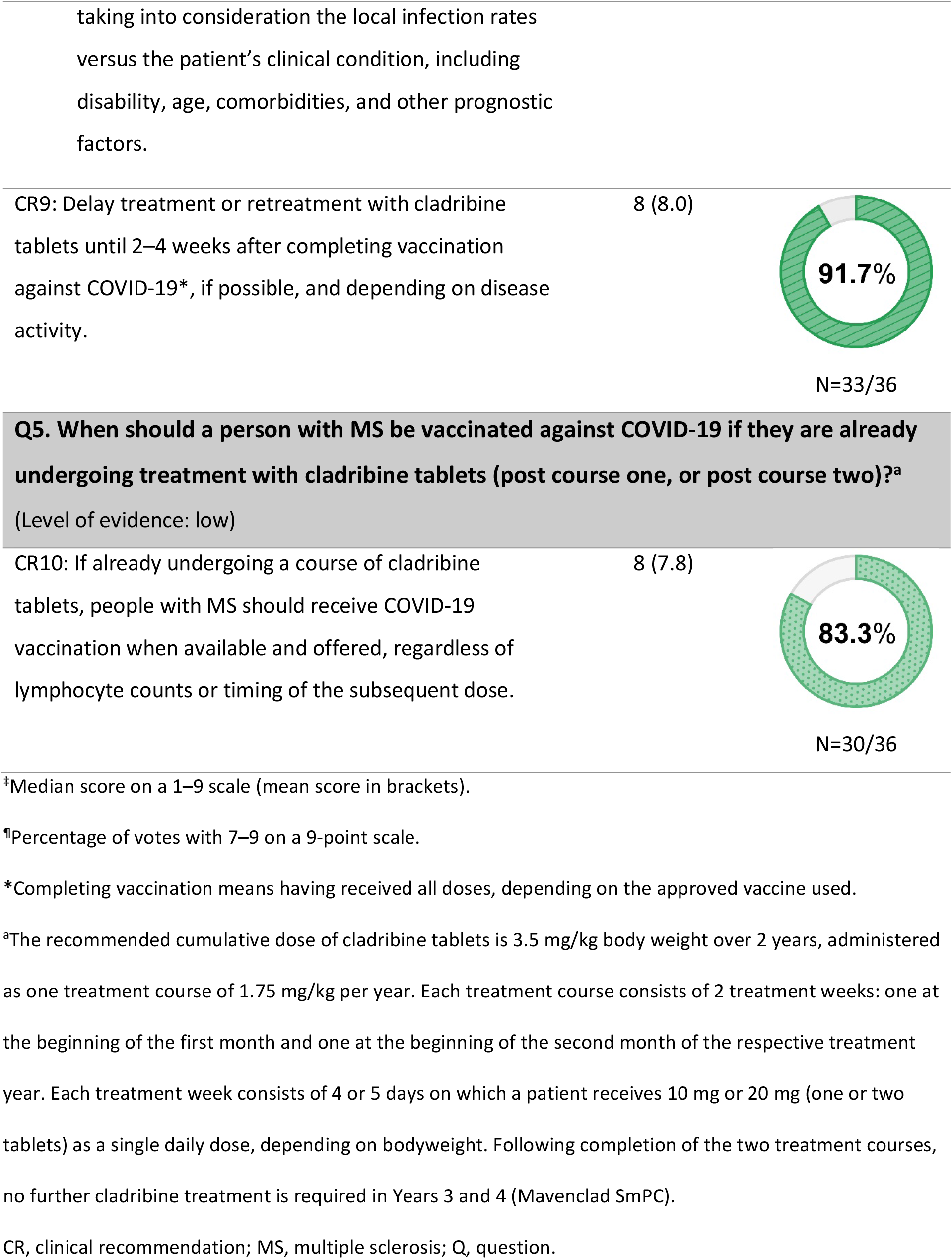
Consensus recommendations to address clinical questions on timing and efficacy of COVID-19 vaccination.

#### Question 4. When should a person with MS be vaccinated against COVID-19 if they are about to start treatment with cladribine tablets (first course in Year 1 or second course in Year 2)?

In the context of vaccination timing, it is important to understand the unique posology of cladribine tablets. The recommended cumulative dose of cladribine tablets is 3.5 mg/kg body weight over 2 years, administered as one treatment course of 1.75 mg/kg per year. Each treatment course consists of 2 treatment weeks: one at the beginning of the first month and one at the beginning of the second month of the respective treatment year. Each treatment week consists of 4 or 5 days on which a patient receives 10 mg or 20 mg (one or two tablets) as a single daily dose, depending on bodyweight. After completing two treatment courses, no further cladribine treatment is required in Years 3 and 4.^38^ The current label for cladribine tablets states that treatment should not be initiated within 4 to 6 weeks after vaccination with live or attenuated live vaccines, but none of the approved COVID-19 vaccines contain live or attenuated live virus.^38^ There are few data regarding timing of vaccinations before starting cladribine tablets. In the small retrospective MAGNIFY-MS analysis, three patients received vaccination against varicella-zoster virus (VZV) before treatment with cladribine tablets. Seroprotective VZV titers were maintained over 6 months post-initiation with cladribine tablets, despite lymphocyte count reduction.^36^ Clinical recommendations to address question 4 are provided in Table 2.

#### Question 5. When should a person with MS be vaccinated against COVID-19 if they are already undergoing treatment with cladribine tablets (post-course one, or post-course two)?

There are no controlled clinical studies specifically investigating vaccine responses in people with MS receiving cladribine tablets. There are data from the small number of patients in the MAGNIFY-MS and CLOCK-MS trials, described earlier.^35,36^ A positive COVID-19 serology test was reported for 17/17 patients with MS receiving cladribine tablets, but the timing around infection and dosing with cladribine tablets in these patients is unknown.^32^ As described earlier, data on the immune-cell responses following treatment with cladribine tablets may have relevance to the timing of vaccinations.^37^ The National Multiple Sclerosis Society recently advised that currently available limited data do not suggest that timing of the vaccine in relation to cladribine tablets dosing is likely to make a significant difference in vaccine response,^16^ while other national guidelines suggest to wait around 3 months before administering the vaccine.^15^ The clinical recommendation to address question 5 is provided in Table 2.

### Safety

#### Question 6. Based on currently available data, are the COVID-19 vaccines safe for use in people with MS receiving cladribine tablets?

There is a high level of evidence in the literature for the safety and tolerability of COVID-19 vaccines in the general population. In Phase 3 randomized controlled trials, vaccines (ChAdOx1 nCoV-19, Gam-COVID-Vac [Sputnik V], mRNA-1273, and BNT162b2) were generally well tolerated and demonstrated acceptable safety profiles.^25-28^ However, at the time of voting, there was no available evidence investigating the safety of COVID-19 vaccination in patients with MS, nor specific analyses looking at patients on cladribine tablets. Clinical recommendations to address question 6 are provided in Table 3.

**Table 3:**
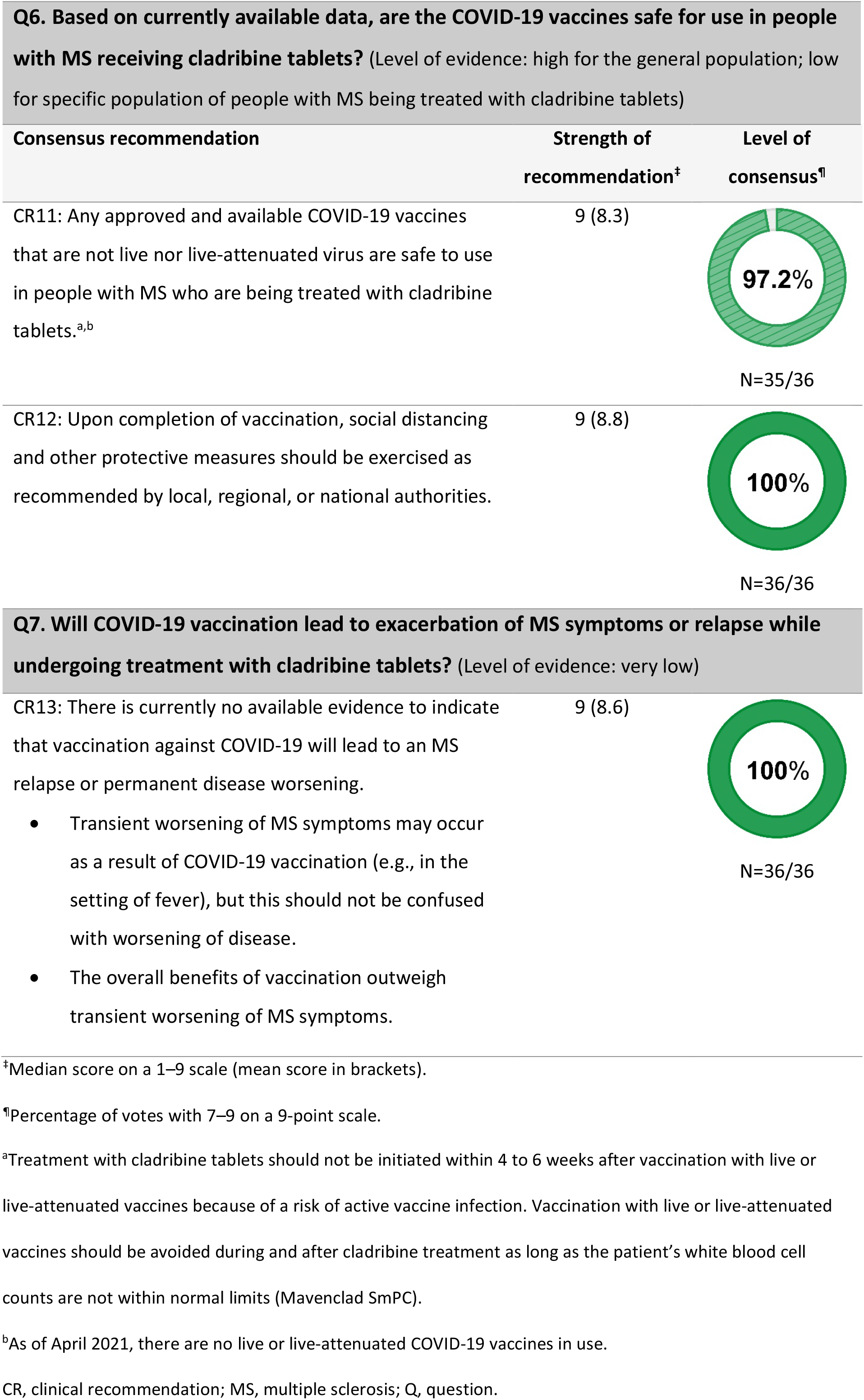
Consensus recommendations to address clinical questions on safety of COVID-19 vaccination.

#### Question 7. Will COVID-19 vaccination lead to exacerbation of MS symptoms or relapse while undergoing treatment with cladribine tablets?

Recent (as of 5 March 2021) global advice from the MS International Federation for the general MS population suggest that “*there is no evidence that people with MS are at higher risk of complications from the mRNA, non-replicating viral vector, inactivated virus or protein COVID-19 vaccines, compared to the general population”*, and that these types of vaccine *“are not likely to trigger an MS relapse or to worsen chronic MS symptoms*.*”*^12^ The clinical recommendation to address question 7 is provided in Table 3.

## Discussion

The clinical recommendations described here provide practical advice on COVID-19 vaccination for people with MS treated with cladribine tablets. The complete faculty of 38 MS experts from 19 countries voted on the recommendations, lending a quality and strength to their relevance. All but one recommendation achieved 90–100% consensus among the faculty.

The recommendation achieving the lowest level of consensus was around the timing of vaccination in patients receiving cladribine tablets: CR10 “*If already undergoing a course of cladribine tablets, people with MS should receive COVID-19 vaccination when available and offered, regardless of lymphocyte counts or timing of the subsequent dose*.*”* The reasons of some faculty members for not strongly agreeing with this statement are provided in Supplementary Table 3. Three of the EF experts commented that the timing of vaccination would depend on the level of lymphocytes or reconstitution. This is a topic of debate in the MS community because most DMTs reduce immune cells, including lymphocytes. Nevertheless, it is important to note that this recommendation still met the criteria for consensus. At present, there is no absolute total lymphocyte count that predicts an antibody response to a vaccine or, for that matter, the outcome of COVID-19 infection.^33, 39-42^ Reasons for this are complex and may relate to the fact that circulating lymphocytes represent only 2–3% of the total lymphocyte pool and do not reflect differences within the T-cell pool; for example, comparing the potential naïve and responsive T cells.^43, 44^ In addition, functional factors, such as immunosenescence, are not reflected in the total lymphocyte counts.^44, 45^ Other relevant factors that predict an antibody response are relevant to the lymph node or secondary lymphoid organ function, in particular the integrity and functioning of germinal centres.^44, 46^

An important topic of discussion among the SC members was whether to include the need for a COVID-19 antibody titer measurement following vaccination. The decision was not to recommend such measurements. It is the opinion of the SC members that there could be some level of protection after COVID-19 vaccination in the absence of a robust humoral B-cell response, mediated by T cells, but these responses are more difficult to measure and standardize. In general, age is an important factor related to antibody production. In people with MS, older patients were at a greater risk of serious COVID-19, including those without comorbidities.^29–31, 33^ Lastly, another complicating factor is the need to standardize testing, since different test assays may report different antibody response results.^47, 48^

One of the weaknesses of the consensus was the limited availability of data; that is, specific studies looking at SARS-CoV-2 antibody responses or COVID-19 vaccination in patients with MS. Large, controlled trials on vaccination are needed, particularly with respect to people with MS on DMTs. In the time since voting took place, new evidence has been published in these areas.^49-51^ Published data from these new studies lend support to at least some of the recommendations described here.

The efficacy of humoral response to the BNT162b2 COVID-19 vaccine was assessed in 125 people with MS; of these, 23 were receiving treatment with cladribine tablets (median time from last treatment dose to vaccination was 7.1 months). All patients treated with cladribine tablets were efficiently vaccinated and developed a protective SARS-CoV-2 antibody titer, even with vaccination as early as 4.4 months after the last treatment dose.^50^ It is reassuring to see that people with MS receiving cladribine tablets mounted good immune responses to COVID-19 vaccination, but it is important to note that the sample size in this study was small, and additional trials with a larger number of patients are needed. It should be noted that, at the time of this consensus exercise, no data on the clinical effectiveness of the COVID-19 vaccines were available from people with MS, whether treated with DMTs or not.

Further recently published data of interest regarding efficacy includes analysis of SARS-CoV-2 antibodies in a large cohort of MS patients from Amsterdam.^51^ Of 546 patients, 64 (11.7%) were positive for SARS-CoV-2 antibodies. Within this cohort, 74.2% of patients were receiving DMTs, and SARS-CoV-2 antibodies were less prevalent in patients using injectable drugs and in patients treated with ocrelizumab.^51^

The safety of the BNT162b2 COVID-19 vaccine was assessed in a cohort of adults with MS: 555 patients received the first dose and 435 received the second dose, of which 414 (74.6%) and 326 (74.9%) were being treated with immunomodulatory drugs, respectively. The most common reported adverse events were local pain at the injection site, fatigue, headaches, muscle or joint pain, and flu-like symptoms manifested as fever, chills, or a combination of both. No increased risk of relapse activity was noted over a median follow-up of 20 and 38 days after first and second vaccine doses, respectively. The rate of patients with acute relapse was 2.1% and 1.6% following the first and second doses, respectively, which is similar to the rate in non-vaccinating patients during the corresponding period.^49^ It is encouraging that in this study, people with MS did not incur more frequent or severe adverse events in response to COVID-19 vaccination compared with the general population, but additional studies with more patients are required.

Further recent evidence that may be of interest is outlined in Supplementary Table S5.

## Conclusion

These consensus-based recommendations represent the opinions and perspectives of a group of international experts within the field of MS. A thorough review of the literature was performed, and the experts carefully considered recent research when formulating their responses. The responses to the questions posed in this expert opinion program are expected to cover the most significant and regularly addressed aspects of COVID-19 vaccination for patients with MS taking cladribine tablets.

These recommendations were developed to provide timely guidance in the absence of much published data on COVID-19 vaccination in people with MS receiving cladribine tablets, and to reflect the knowledge, evidence, and opinion in 2021. Updates to these recommendations will be made as new evidence emerges. This expert opinion is expected to be a concise and thorough resource for medical professionals, with the aim of assisting them in making informed decisions regarding COVID-19 vaccinations in this group of patients to improve their standard of care.

## Data Availability

Submission in progress

## Acknowledgements

Emma East and Stephanie Wasek of Bedrock Healthcare Communications provided editorial assistance, funded by Merck KGaA (Darmstadt, Germany).

The SC would like to thank all the experts who contributed their knowledge to this program by voting on the draft recommendations. Listed below are 27 EF members who are happy to be acknowledged in this document:

Aaron Boster^1^, Wallace Brownlee^2^, Elisabeth G. Celius^3^, Marinella Clerico^4^, Dominique Dive^5^, Juha-Pekka Erälinna^6^, Mark Freedman^7^, Claudio Gasperini^8^, Nikolaos Grigoriadis^9^, Eva Kubala Havrdová^10^, Barry Hendin^11^, Dimitrios Karussis^12^, Natalia Khachanova^13^, Joanna Kitley^14^, Barbara Kornek^15^, Melinda Magyari^16^, Mathias Mäurer^17^, Virginia Meca-Lallana^18^, Marcello Moccia^19^, Ester Moral^20^, David Paling^21^, Marco Salvetti^22^, Klaus Schmierer^23^, Aksel Siva^24^, Maria-Pia Sormani^25^, Natalia Totolyan^26^, Bart Van Wijmeersch^27^

The Boster Center for Multiple Sclerosis, Columbus, Ohio^1^, National Hospital for Neurology and Neurosurgery, Queen Square, London, UK^2^, Department of Neurology, Oslo University Hospital, Oslo, Norway^3^, Clinical and Biological Sciences Department, University of Torino, Torino (IT)^4^, University Hospital of Liège, Liège, Belgium^5^, Mehiläinen NEO, Turku, Finland^6^, University of Ottawa, Department of Medicine,, the Ottawa Hospital Research Institute, Ottawa, Ontario, Canada^7^, Department of Neuroscience, San Camillo-Forlanini Hospital, Rome, Italy^8^, B’ Department of Neurology, Aristotle University of Thessaloniki, Thessaloniki, Greece^9^, Department of Neurology and Center for Clinical Neuroscience, First Medical Faculty, Charles University, Prague, Czech Republic^10^, University of Arizona, Phoenix Arizona, USA^11^, Hadassah-Hebrew University Medical Organization, Jerusalem, Israel^12^, Pirogov Russian National Research Medical University, Moscow, Russia^13^, Wessex Neurological Centre, Southampton, England^14^, Medical University of Vienna, Department of Neurology, Vienna, Austria^15^, Danish Multiple Sclerosis Centre, Copenhagen University Hospital, Rigshospitalet, Copenhagen, Denmark^16^, Klinikum Würzburg Mitte, Würzburg, Germany^17^, Hospital Universitario de la Princesa, Madrid, Spain^18^, University of Naples, Naples, Italy^19^, Moises Broggi Hospital, Barcelona, Spain^20^, Royal Hallamshire Hospital, Sheffield, UK^21^, Sapinza University, Rome, Italy^22^, Queen Mary University of London, London, UK^23^, Istanbul University Cerrahpasa School of Medicine, Istanbul, Turkey^24^, Department of Health Sciences, University of Genoa, Italy^25^, Department of Neurology, 1st Saint Petersburg Pavlov State Medical University, Saint Petersburg, Russia^26^, Universitair MS Centrum Hasselt - Pelt, Belgium, Hasselt-Pelt, Belgium^27^

