## Supplemental tables S1 to S5 for "Expert Opinion on COVID-19 Vaccination and the Use of Cladribine Tablets in Clinical Practice"

**Supplementary Table S1. Further reading on the consensus themes**

| Authors | Title | Publication and date | Article type |
|---|---|---|---|
| *Patient selection* | | | |
| Giovannoni G, Hawkes C, Lechner-Scott J, et al. | The COVID-19 pandemic and the use of MS disease-modifying therapies. | *Mult Scler Relat Disord* 2020; 39: 102073. | Commentary |
| Hartung HP and Aktas O. | COVID-19 and management of neuroimmunological disorders. | *Nat Rev Neurol* 2020; 16(7): 347–348. | Commentary |
| Jack D, Nolting A and Galazka A. | Favorable outcomes after COVID-19 infection in multiple sclerosis patients treated with cladribine tablets. | *Mult Scler Relat Disord* 2020; 46: 102469. | Correspondence |
| *Timing* | | | |
| No authors listed | Cladribine for multiple sclerosis. | *Drug Ther Bull* 2018; 56(2): 21–24. DOI: 10.1136/dtb.2018.2.0590. | Review |
| Meca-Lallana V, Aguirre C, Cardeñoso L, et al. | Establishment of a safety protocol for the administration of treatments in multiple sclerosis during the SARS-CoV-2 pandemic. | *Mult Scler Relat Disord* 2020; 44: 102244. | Correspondence |
| Preziosa P, Rocca MA, Nozzolillo A, et al. | COVID-19 in cladribine-treated relapsing-remitting multiple sclerosis patients: A monocentric experience. | *J Neurol* 2020; 268: 2697–2699. | Case report (N=2) |
| Rohit B, Padma Srivastava MV, Khurana D, et al. | Consensus statement on immune modulation in multiple sclerosis and related disorders during the COVID-19 pandemic: Expert group on behalf of the Indian Academy of Neurology. | *Ann Indian Acad Neurol* 2020; 23(1): S5. | Consensus |

| | | | |
|---|---|---|---|
| Sellner J and Rommer PS. | Multiple sclerosis and SARS-CoV-2 vaccination: Considerations for immune-depleting therapies. | *Vaccines* 2021; 9(2); 99. | Review |
| Zheng C, Kar I, Chen CK, et al. | Multiple sclerosis disease-modifying therapy and the COVID-19 pandemic: Implications on the risk of infection and future vaccination. | *CNS Drugs* 2020; 34(9): 879–896. | Current opinion |
| Al Jumah M, Abulaban A, Aggad H, et al. | Managing multiple sclerosis in the COVID-19 era: A review of the literature and consensus report from a panel of experts in Saudi Arabia. | *Mult Scler Relat Disord* 2021; 51: 1–8. DOI: 10.1016/j.msard.2021.102925. | Review and recommendations |
| Inshasi J, Alroughani R, Al-Asmi A, et al. | Expert consensus and narrative review on the management of multiple sclerosis in the Arabian Gulf in the COVID-19 era: Focus on disease-modifying therapies and vaccination against COVID-19. | *Neurol Ther* 2021; 1–17. DOI: 10.1007/s40120-021-00260-5. | Expert consensus and narrative review |
| Inshasi JS, Alfahad S, Alsaadi T, et al. | Position of cladribine tablets in the management of relapsing-remitting multiple sclerosis: An expert narrative review from the United Arab Emirates. | *Neurol Ther* 2021; 1–20. DOI: 10.1007/s40120-021-00243-6. | Expert narrative review |
| *Efficacy* | | | |
| Apóstolos-Pereira SL, Silva GD, Disserol CCD, et al. | Management of central nervous system demyelinating diseases during the coronavirus disease 2019 pandemic: A practical approach. | *Arq Neuropsiquiatr* 2020; 78(7): 430–439. DOI: 10.1590/0004-282X20200056. | Review |
| Celius EG. | Normal antibody response after COVID-19 during treatment with cladribine. | *Mult Scler Relat Disord* 2020; 46: 102476. | Commentary |

| | | | |
|---|---|---|---|
| Ciotti JR, Valtcheva MV, Cross AH. | Effects of MS disease-modifying therapies on responses to vaccinations: A review | *Mult Scler Relat Disord* 2020; 102439. | Review |
| De Angelis M, Petracca M, Lanzillo R, et al. | Mild or no COVID-19 symptoms in cladribine-treated multiple sclerosis: Two cases and implications for clinical practice. | *Mult Scler Relat Disord* 2020; 45:102452. | Correspondence (N=2) |
| Bsteh G, Assar H, Hegen H, et al. | COVID-19 severity and mortality in multiple sclerosis are not associated with immunotherapy: Insights from a nation-wide Austrian registry. | *PLoS One* 2021; 16(7): e0255316. DOI: 10.1371/journal.pone.0255316. eCollection 2021. | Population-based study (N=126) |
| Buttari F, Bruno A, Dolcetti E, et al. | COVID-19 vaccines in multiple sclerosis treated with cladribine or ocrelizumab. | *Mult Scler Relat Disord* 2021; 52: 102983. DOI: 10.1016/j.msard.2021.102983. | Correspondence |
| *Safety* | | | |
| Gelibter S, Orrico M, Filippi M, et al. | COVID-19 with no antibody response in a multiple sclerosis patient treated with cladribine: Implication for vaccination program? | *Mult Scler Relat Disord* 2021; 49: 102775. | Correspondence |
| Margoni M , Annovazzi P, Prosperini L, et al. | A multicentre, real-life study on the risk of lymphopenia and infections discloses a favourable safety profile of cladribine in MS patients. | *Mult Scler J* 2020; 26(S3): 252–252. | Observational multicenter study (N=236) |
| Achiron A, Dolev M, Menascu S, et al. | COVID-19 vaccination in patients with multiple sclerosis: What we have learnt by February 2021. | *Mult Scler J* 2021; 27(6): 864–870. DOI: 10.1177/13524585211003476. | Observational study (N=574) |
| Achiron A, Mandel M, Dreyer-Alster S, et al. | Humoral immune response to COVID-19 mRNA vaccine in patients with multiple sclerosis treated with high-efficacy disease-modifying therapies. | *Ther Adv Neurol Disord* 2021; 14: 1–8. DOI: 10.1177/17562864211012835. | Observational cohort study (N=172) |
| *Multiple themes/general guidance* | | | |
| Kelly H, Sokola B, Abboud H. | Safety and efficacy of COVID-19 vaccines in multiple sclerosis patients | *J Neuroimmunol* 2021; 356: 577599. DOI: | Review |

| | | | |
|---|---|---|---|
| | | 10.1016/j.jneuroim.2021.577599. | |
| Nesbitt C, Rath L, Zhong M, et al. | Vaccinations in patients with multiple sclerosis: Review and recommendations. | *Med J Aust* 2021; 214(8); 350–354. DOI: 10.5694/mja2.51012. | Review and recommendations |
| Woopen C, Schleußner K, Akgün K, et al. | Approach to SARS-CoV-2 vaccination in patients with multiple sclerosis. | *Front Immunol* 2021; 12: 2458. DOI: 10.3389/fimmu.2021.701752. | Perspective |

COVID-19, coronavirus disease 2019; SARS-CoV-2, severe acute respiratory syndrome coronavirus-2.

**Supplementary Table S2. Patient selection—verbatim reasons provided for scoring 6 or less on a clinical recommendation**

| **Q1. Should all people with MS receiving cladribine tablets be vaccinated against COVID-19?** (Level of evidence: high for the general population; low for specific population of people with MS being treated with cladribine tablets) |
|---|
| CR2: In general, people with MS are not more likely to contract COVID-19 or to experience a more-severe COVID-19 course. However, limited data suggest that patients on some MS therapies are at risk of more severe COVID-19 outcomes.<br>• Early data suggest that people with MS receiving cladribine tablets are generally not at greater risk of serious disease and/or a severe outcome from COVID-19 compared with the general population and other people with MS who acquired COVID-19.<br><br>**Reasons for voting 6 or less:**<br>*“I strongly agree with "Early data suggest that people with MS receiving cladribine tablets are generally not at greater risk of serious disease and/or a severe outcome from COVID-19 compared with the general population, and other people with MS who acquired COVID-19".*<br>*However, I partly agree with "In general, people with MS are not more likely to contract COVID-19 or to experience a more-severe COVID-19 course; however, limited data suggest that patients on some MS therapies are at-risk of more severe COVID-19 outcomes." This sentence is based on real-world evidence (selection bias - patients who die from COVID19 cannot attend the MS centre to* |

| |
|---|
| *report on their death), while it should be coming from population-based studies. Also, people with MS are more at risk of infection and worse outcomes, so it is still counterintuitive why this should not be the case for COVID19. Finally, this sentence is not in line with Q2 (Should people with MS treated with cladribine tablets be prioritized for COVID-19 vaccination? Level of evidence: high (risk-factor subgroups are well-reported in the literature)).”*<br>*“The number of patients treated with cladribine in the several cohort published is small”* |
| CR3: People with MS receiving treatment with cladribine tablets who have already experienced infection with SARS-CoV-2 should still be vaccinated, according to their national guidelines, after resolution of COVID-19 symptoms.<br><br>**Reasons for voting 6 or less:**<br>*“It depends on the level of IgG”*<br>*“It is not clear if and when people who had Covid in general need to be vaccinated”*<br>*“Should be vaccinated or not - depends on i) time after COVID19 resolution; ii) anti-CoVID-19 Ab status - if these statements are not provided by national guidelines for vaccination”* |
| **Q2. Should people with MS treated with cladribine tablets be prioritized for COVID-19 vaccination?** Level of evidence: high (risk-factor subgroups are well-reported in the literature) |
| CR4: People with MS should be prioritized for COVID-19 vaccination on an individual basis depending on risk factors, including disability, age, comorbidities that increase the risk for severe COVID-19 course, and whether they are about to start treatment with cladribine tablets.<br><br>**Reasons for voting 6 or less:**<br>*“People with MS need to access healthcare frequently e.g. for consultations, blood tests, MRI scans etc. All people with MS should be prioritized for vaccination. This is the advice in the UK.”* |

Consensus on CR1 achieved 100%, hence is not included in this table.

COVID-19, coronavirus disease 2019; CR, consensus recommendation; IgG, immunoglobulin G; MRI, magnetic resonance imaging; MS, multiple sclerosis; UK, United Kingdom.

**Supplementary Table S3. Timing and efficacy—verbatim reasons provided for scoring 6 or less on a clinical recommendation**

| **Q3. Should all people with MS on cladribine tablets receive a vaccination against COVID-19 at the time it is offered?** (Level of evidence: moderate) |
|---|
| CR6: Treatment with cladribine tablets is unlikely to have a major impact on vaccine responses.<br>• Some degree of immunity is preferable to no immunity in individuals who may have a reduced response to a vaccine.<br><br>**Reasons for voting 6 or less**<br>*"I don't feel that there is enough evidence to say that cladribine is unlikely to have an impact on vaccine effectiveness, but the second point is definitely true, and vaccination should be given."*<br>*"..Immune depletion is not always mild"*<br>Reason not provided for 1 vote of score 6 |
| **Q4. When should a person with MS be vaccinated against COVID-19 if they are about to start treatment with cladribine tablets (first course in Year 1 or second course in Year 2)?** (Level of evidence: low) |
| CR8: In general, increased risk of MS disease worsening is the greater threat to long-term health in people with MS than SARS-CoV-2 infection. As a result, treatment initiation with cladribine tablets and long-term stability of MS should be prioritized over vaccination.<br>• The risk of COVID-19 should be balanced against the benefit of treatment with cladribine tablets, taking into consideration the local infection rates versus the patient's clinical condition, including disability, age, comorbidities, and other prognostic factors.<br><br>**Reasons for voting 6 or less**<br>*"There is no evidence that such a delay affects consequent immune response"*<br>*"Looking at vaccine responses, a delay of 10-14 days would suffice"*<br>*"Delay in treatment could adversely affect the MS. Currently COVID dosing is 4 months apart in our country [so] at least the 2nd dose will be done at a better time relative to cladribine treatment"* |
| CR9: Delay treatment or retreatment with cladribine tablets until 2–4 weeks after completing vaccination against COVID-19, if possible, and depending on disease activity.<br><br>**Reasons for voting 6 or less**<br>*"I believe the priority is herd immunity. Of course, one might consider delaying COVID19 vaccination, if there is the possibility caregivers/close contacts are vaccinated."* |

*“Individual decision, depending on the expected timing of immunisation”*

*“Potential though not accurate prognostic factors for MS are established with regard to long-term disability and overall deterioration of patient's health status. However, this is not yet the case for COVID-19 outcome.”*

**Q5. When should a person with MS be vaccinated against COVID-19 if they are already undergoing treatment with cladribine tablets (post-course one, or post-course two)?**

(Level of evidence: low)

CR10: If already undergoing a course of cladribine tablets, people with MS should receive COVID-19 vaccination when available and offered, regardless of lymphocyte counts or timing of the subsequent dose.

**Reasons for voting 6 or less**

*“The level of lymphocytes is important”*

*"This recommendation would apply to vaccines taken 4.5 months after last CT dosing (as per Achiron et al data). We don’t have enough evidence to support such recommendation during the first 3 months, at the peak of the lymphopenia, especially in the few patients that develop Grade III lymphopenia. In the CLOCK study the only patient with GIII lymphopenia at the time of vaccination did not double its antibody titres to Influenza. The only exception would be if the patient loses his/her turn to vaccinate in case of delay"*

*“Slightly ambiguously worded question. If pt is in between courses then I strongly agree. If 2nd course is due, I would delay slightly to facilitate vaccination first”*

*“Ideally, one could postpone the 2nd course for up to 6 months to give a 4-6 week window to allow for the vaccine response”*

*“To have good vaccination response it would be better to wait for at least partial immune reconstitution, e.g. at least 3 months.”*

*“I would advise the same time lapse as for 1st treatment cycle (2-4 weeks), simply to be able to separate adverse effects.”*

Consensus on CR5 and CR7 achieved 100%, hence are not included in this table.

COVID-19, coronavirus disease 2019; CR, consensus recommendation; CT, cladribine tablets; MS, multiple sclerosis; SARS-CoV-2, severe acute respiratory syndrome coronavirus-2.

**Supplementary Table S4. Safety—reasons provided for scoring 6 or less on a clinical recommendation**

| **Q6. Based on currently available data, are the COVID-19 vaccines safe for use in people with MS receiving cladribine tablets?** (Level of evidence: high for the general population; low for specific population of people with MS being treated with cladribine tablets) |
|---|
| CR11: Any approved and available COVID-19 vaccines that are not live nor live-attenuated virus are safe to use in people with MS who are being treated with cladribine tablets. |
| **Reasons for voting 6 or less** |
| *"Avoid all live vaccines."* |

Consensus on CR12 and CR13 achieved 100%, hence are not included in this table.

COVID-19, coronavirus disease 2019; CR, consensus recommendation; MS, multiple sclerosis.

**Supplementary Table S5. Existing professional and patient society guidance at the time of creating clinical recommendations**

| Region | Society | Relevant original passages considered |
|---|---|---|
| International | MS International Federation[1] | • It is safe to receive a COVID-19 vaccine when you are on MS DMTs. Delaying the start of a DMT, or altering DMT timing, is not a safety issue—it is a strategy to allow the vaccine to be fully effective.<br>• Some DMTs may reduce the effectiveness of the COVID-19 vaccinations.<br>• Even once you have received the vaccine, it is important to continue to take precautions against COVID-19.<br>• If you are about to start cladribine, consider getting fully vaccinated* 2–4 weeks before starting cladribine. If you are already taking cladribine, the currently available limited data does not suggest that timing the vaccine in relation to your cladribine dosing is likely to make a significant difference in vaccine response. Getting the vaccine when it becomes available to you may be more important than coordinating timing of the vaccine with your cladribine treatment. If you are due for your next treatment course, when possible, resume cladribine 2–4 weeks after getting fully vaccinated*. |

| | | |
|---|---|---|
| | | *Fully vaccinated = once you have received the single dose of the J&J vaccine or the second dose of any other type of vaccine. |
| Canada | Multiple Sclerosis Society of Canada[2] | • The effectiveness of COVID-19 vaccination in people with MS and DMT is thus far unknown.<br>• Based on data from previous studies of other vaccines and DMTs, getting the COVID-19 vaccine while on any DMT is safe.<br>• Some DMTs may make the vaccine less effective but it will still provide some protection. For those taking Kesimpta, Lemtrada, Mavenclad, Ocrevus, or Rituxan (rituximab) — you may need to coordinate the timing of your vaccine with the timing of your DMT dose. The decision of when to get the COVID-19 vaccine should include an evaluation of your risk of COVID-19, including your occupation, and the current state of your MS. Work with your MS healthcare provider to determine the best schedule for you. |
| Germany | Deutsche multiple sclerosis Gesellschaft Bundesverband (Dmsg)[3] | • Cladribine (Mavenclad): No data on vaccination studies are available yet. Due to the mechanism of action, a reduced vaccination response is to be expected at least temporarily, in the first 6 months after the therapy cycle. |
| United Kingdom | Association of British Neurologists[4] | • All COVID-19 vaccines are safe for neurology patients. Take up whichever vaccination you are offered without delay regardless of your age, state of health or medication that you're receiving, unless you have a specific contraindication such as severe allergy. None of the COVID-19 vaccines are 'live' and therefore cannot cause infection. |
| United Kingdom | MS Society[5,6] | • There may be a reduced response to a vaccine with cladribine (Mavenclad), and it may be advisable to wait 3 months after a course before having a vaccine.<br>• A second course of cladribine is usually given after 12 months, but this can safely be delayed for several months without concerns for a return of disease activity, allowing some flexibility to schedule vaccination in advance.<br>• If starting for the first time, you should discuss with your team whether it might be preferable to wait until you have been vaccinated. The risks of this strategy will depend on your individual case and on the availability of the vaccine. |
| United States | American Academy of Neurology (AAN)[7] | • The AAN supports the vaccination recommendations by the FDA.<br>• In order to mitigate the spread of COVID-19 to the high-risk patients who seek our care, the AAN |

| | | |
|---|---|---|
| | | strongly encourages that all eligible neurology healthcare providers become vaccinated against COVID-19 and support vaccination for all patients who qualify. In addition, the AAN strongly supports efforts to ensure all patients have equitable access to COVID-19 vaccination. |
| United States | National Multiple Sclerosis Society[8] | • If you are about to start Mavenclad, consider getting fully vaccinated* 2–4 weeks before starting Mavenclad.<br>• If you are already taking Mavenclad, the currently available limited data does not suggest that timing of the vaccine in relation to your Mavenclad dosing is likely to make a significant difference in vaccine response. Getting the vaccine when it becomes available to you may be more important than coordinating timing of the vaccine with your Mavenclad treatment. If you are due for your next treatment course, when possible, resume Mavenclad 2–4 weeks after getting fully vaccinated*. Work with your MS healthcare provider to determine the best schedule for you.<br>*Fully vaccinated = two doses of the mRNA (Pfizer BioNTech or Moderna) or one dose of the vector vaccine (J&J) |

AAN, American Academy of Neurology; COVID-19, coronavirus disease 2019; DMT, disease-modifying therapy; FDA, Food and Drug Administration; J&J, Johnson and Johnson; mRNA, messenger ribonucleic acid; MS, multiple sclerosis.


1. MS International Federation. Global COVID-19 advice for people with MS, https://www.msif.org/news/2020/02/10/the-coronavirus-and-ms-what-you-need-to-know/ (2021, accessed May 23, 2021).
2. Multiple Sclerosis Society of Canada. COVID-19 vaccine guidance for people living with MS, https://mssociety.ca/resources/news/article/covid-19-vaccine-guidance-for-people-living-with-ms (2021, accessed May 23, 2021).
3. Deutsche multiple sclerosis Gesellschaft Bundesverband (Dmsg). Impfungen und multiple sklerose, https://www.dmsg.de/corona-virus-und-ms/impfung0/ (2021, accessed May 23, 2021). [German]
4. Association of British Neurologists. Guidance on vaccination for COVID-19 and neurological conditions, https://cdn.ymaws.com/www.theabn.org/resource/collection/65C334C7-30FA-45DB-93AA-74B3A3A20293/ABN_Guidance_on_COVID-19_Vaccinations_for_people_with_neurological_conditions_9.1.21.pdf (2021, accessed August 6, 2021)
5. MS Society. COVID-19 coronavirus and MS treatments, https://www.mssociety.org.uk/about-ms/treatments-and-therapies/disease-modifying-therapies/covid-19-coronavirus-and-ms (2021, accessed May 14, 2021).
6. MS Society. MS Society Medical Advisers consensus statement on MS treatments and COVID-19 vaccines, https://www.mssociety.org.uk/what-we-do/news/ms-society-medical-advisers-release-consensus-statement-covid-19-vaccines (2021, accessed May 23, 2021).
7. American Academy of Neurology (AAN). Position statement: COVID-19 vaccination, https://www.aan.com/siteassets/home-page/policy-and-guidelines/policy/position-

statements/21-positionstatement-covid-19-vaccination-v001.pdf%20, (2021, accessed May 23, 2021).
8. National Multiple Sclerosis Society. Timing MS medications with COVID-19 vaccines, https://www.nationalmssociety.org/coronavirus-covid-19-information/multiple-sclerosis-and-coronavirus/covid-19-vaccine-guidance/Timing-MS-Medications-with-COVID-19-Vaccines (2021, accessed May 14, 2021).
